# A multimodal protocol for assessing real-world monitoring of lower-limb prosthesis use

**DOI:** 10.64898/2026.08.21.26360982

**Authors:** Mustafa Ahmed, Sophia Otálora, Sauvik Das Gupta, Gaku Kutsuzawa, Abdullah Akaydin, Julien Le Kernec, Yoshiyuki Kobayashi, Encarna Micó-Amigo

## Abstract

Prosthesis non-use and abandonment remain common among people with lower-limb amputation, yet current outcome measures capture only limited aspects of how prostheses are used in everyday life. Clinical assessments are typically conducted in controlled settings and rely on self-report or aggregate activity counts, which do not adequately represent functional performance, physiological effort, or lived experience during real-world prosthesis use. Wearable and ambient sensing offer a means of addressing this gap, but existing approaches tend to measure single dimensions in isolation and are rarely validated against laboratory reference standards before free-living deployment. This protocol describes an integrated multimodal framework for assessing real-world lower-limb prosthesis use across three complementary domains: classification of activities of daily living, estimation of energy expenditure, and assessment of emotional state. Approximately 40 adults with unilateral transfemoral or transtibial amputation complete a two-phase protocol. In the laboratory phase, wearable inertial, physiological, and ambient sensing are validated against established reference standards, including video annotation and indirect calorimetry. In the free-living phase, validated models are applied during a single seven-day home monitoring period, unifying all three domains within one deployment. A defined data harmonisation and quality-control procedure aligns heterogeneous sensor streams and preserves traceability between laboratory calibration and free-living measurement, enabling reproducible interpretation of functional behaviour, metabolic cost, and momentary emotional experience in relation to established clinical outcome domains. By integrating multimodal sensing at the level of study design rather than post-hoc analysis, the framework provides a validated, reproducible methodology for characterising prosthesis use beyond the capacity of conventional instruments, offering a transferable approach for real-world monitoring in rehabilitation research.

## Introduction

An estimated 35–40 million people worldwide require prosthetic devices, and this number is expected to increase due to the rising prevalence of chronic conditions such as diabetes and peripheral arterial disease, trauma, and ageing populations [1–3]. In England, major lower-limb amputation affects thousands of individuals each year, with incidence particularly elevated among those with diabetes and vascular disease [4]. When appropriately prescribed, prosthetic Appropriate lower-limb prosthetic provision aims to maximise ambulation, function, and quality of life [5]. Among prosthesis users, self-reported functional mobility and prosthetic use are associated with daily step counts [6], while activity and prosthesis-related factors are associated with community participation [7]. However, despite advances in prosthetic technology and clinical care, prosthesis non-use and abandonment remain important challenges, with approximately 11–22% of lower-limb prosthesis recipients reported to stop using their prosthesis at around one year [5]. This highlights a critical challenge in translating technological provision into sustained functional benefit in everyday life.

Despite substantial advances in prosthetic technologies and clinical provision, the goals of prosthetic care extend beyond restoring locomotion [8, 9]. Effective prosthetic management seeks to support functional independence, maintain quality of life, and enable meaningful participation in activities of daily living (ADL) and broader social roles [10, 11]. The capacity to perform ADL safely and independently is therefore a central outcome of prosthetic rehabilitation, yet how prosthesis users engage with these activities in real-world environments remains poorly characterised [12, 13].

Clinical evaluation of prosthesis use typically relies on structured outcome measures and patient-reported instruments assessing domains such as mobility, physical function, prosthesis experience, and psychosocial adaptation [14]. While these tools provide valuable insights, they are often limited to episodic assessments or retrospective self-report, and may not capture the variability and contextual influences of real-world behaviour, being biased by recall [12, 13]. As a result, how prostheses are actually used in real-world environments remains poorly understood.

Understanding how, how much, where, and for what purpose lower-limb prostheses are used in real-world contexts is essential for informing the design, provision, and evaluation of prosthetic devices and rehabilitation strategies [15]. Real-world behaviour is shaped not only by activity type, but also by environmental context, the use of walking aids, and the user’s physical and emotional state [15, 16]. In particular, walking in people with lower-limb amputation is associated with a higher energy cost than in people without amputation, with the magnitude varying according to amputation level, aetiology, and walking speed [17]. At the same time, psychosocial factors such as confidence, embodiment, and frustration influence how individuals engage with their prosthesis in everyday life [18, 19]. Current assessment methods provide only a limited view of these interacting dimensions, particularly in free-living settings.

Wearable and ambient sensing technologies offer opportunities to address these limitations by enabling continuous and unobtrusive monitoring of behaviour in natural environments [15, 20]. While earlier studies have demonstrated the feasibility of capturing basic metrics such as step count and activity intensity [21, 22], such measures provide limited insight into functional activity patterns and contextual influences [12, 23]. Advances in inertial measurement units, smartwatch-based sensing, and environmental sensors now enable more detailed characterisation of movement and interaction with the environment [24, 25]. However, the integration and complementary roles of these sensing modalities remain insufficiently defined, particularly in prosthetic populations where movement patterns are highly variable [24, 26] and influenced by internal factors, such as residual limb condition, pain, fatigue, muscle strength, balance confidence, and psychosocial state [18, 19], as well as external factors, such as prosthetic fit and alignment, walking-aid use, terrain, environmental barriers, and task-specific demands [27, 28].

In response to these challenges, the present article describes a structured multimodal protocol designed to assess real-world prosthesis use and associated functional outcomes. The protocol integrates wearable and ambient sensing to capture multiple dimensions of daily functioning, including physical activity, metabolic cost, and psychosocial experience. To support these objectives, the protocol combines controlled laboratory assessments for high-fidelity reference data used in validation procedures, and a reduced, low-burden wearable configuration for free-living deployment. Specifically, the objectives of this protocol are to:

1. Co-design the protocol through structured Patient and Public Involvement with lower-limb prosthesis users, clinicians, and industrial partners, ensuring that monitored domains, sensing configurations, and assessment procedures reflect the functional priorities and lived experience of the target population;
2. Establish and validate laboratory-based procedures for wearable-based ADL classification, wearable-derived energy expenditure calibration referenced to indirect calorimetry, and emoji-based emotional state instrument derivation in lower-limb prosthesis users;
3. Develop and evaluate a unified free-living monitoring framework concurrently deploying all three studies through a novel two-stream ADL classification architecture that combines human-in-the-loop ecological validation with autonomous classification, linking functional activity, wearable-derived energy expenditure, and momentary emotional states in prosthesis users.

By integrating wearable and ambient sensing modalities within a unified, validated, and standardised framework, this protocol aims to advance methodological approaches and inform the development of scalable and transferable monitoring strategies for prosthetic care. In addition to supporting proof-of-concept deployment using a minimal wearable configuration, the protocol defines procedures for generating richly annotated and standardised multimodal datasets that may facilitate future methodological development and cross-study reuse, subject to appropriate governance. Through a co-designed process informed by Patient and Public Involvement (PPI), this work establishes a methodological foundation for subsequent validation studies and the advancement of more personalised, evidence-informed approaches to prosthetic care for individuals living with limb loss.

## Materials and methods

### Study design overview

#### Clinical outcome domains

Table 1 summarises clinical domains commonly assessed in prosthetic outcome instruments and illustrates how these domains are operationalised within established questionnaires. These instruments are widely used to evaluate prosthesis use and functional mobility in individuals with limb loss.

**Table 1.**
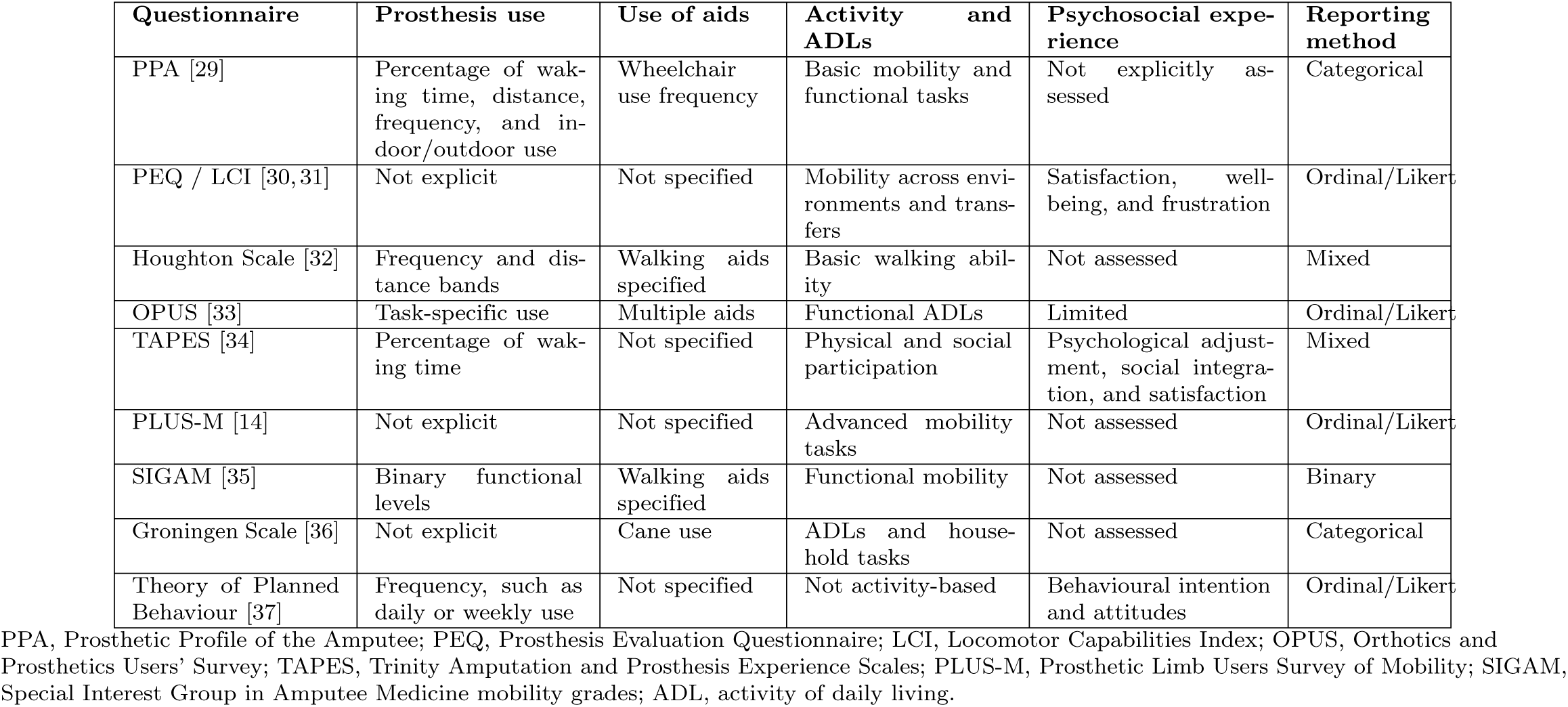
Clinical domains commonly assessed in prosthetic outcome instruments. Reporting methods include binary yes/no responses, categorical responses, ordinal or Likert-type scales, proportional estimates such as percentage or time-based reporting, and mixed formats combining more than one response type.

Across instruments, several recurring constructs are observed. These include patterns of prosthesis use (e.g., frequency and duration of wear, distance travelled, and use across different contexts such as indoor and outdoor mobility), reliance on walking aids during ambulation, and the performance of activities of daily living. Within established outcome measures, these constructs are typically operationalised using time-based metrics (e.g., hours per day, percentage of waking time, days per week), distance categories, task-specific binary responses, and ordinal or Likert-type frequency scales. Reporting formats vary across instruments and are described in the final column of Table 1.

#### Patient and public involvement

PPI was undertaken to inform the design and practical implementation of the proposed monitoring framework. The consultation focused on how domains commonly assessed in established prosthetic outcome instruments (Table 1) could be meaningfully and acceptably captured using multimodal sensing technologies.

Twelve prosthetic users, four clinicians with experience in prosthetic prescription and long-term clinical management, and two industrial partners participated in the consultation process. Participants were presented with a structured overview of the proposed monitoring approach, including laboratory-based ADL assessment, wearable and ambient sensing, treadmill-based physiological testing, and short-term free-living monitoring.

Discussions were organised around functional and experiential domains commonly represented in prosthetic outcome measures, including prosthesis use, mobility-related performance and activities, reliance on walking aids, and lived experience during daily activity. Consultation data were analysed using framework analysis [38], enabling systematic identification and organisation of themes across stakeholder groups. Feedback focused on perceived barriers and limitations to technology-based monitoring, including perceived user burden, acceptability of sensor placement, feasibility of sustained and continuous monitoring, and relevance to everyday prosthesis use.

PPI input contributed to protocol design across three key areas. First, prosthetic user feedback informed the selection of ADL for algorithm development and validation, identified through a combination of established clinical instruments and domains highlighted during consultation; the source of each activity is indicated in Table 4. Second, sensor configurations and placement were refined to minimise monitoring burden and enhance acceptability. Third, the frequency and format of emotional state sampling were adjusted to reduce disruption to daily routines. A detailed account of stakeholder perspectives and their influence on protocol design decisions is provided in the Discussion.

#### Phases of the protocol

The protocol is organised around three complementary studies (classification and monitoring of ADL, energy expenditure estimation and assessment, and emotional states evaluation and contextualisation), with each study progressing from laboratory-based validation to free-living implementation. Table 2 provides an overview of instrumentation, procedures, and analyses across both phases.

**Table 2.** Study framework overview: instrumentation, procedures, and data analysis by study and phase.

| Study | Phase | Instrumentation | Procedures | Data analysis |
| --- | --- | --- | --- | --- |
| ADL study | Phase 1: Lab validation | *Gold standard:* video cameras. *Wearables:* smartwatch with ADL Recorder app; Axivity AX6 on the lower back; six Xsens IMUs on the sternum, anterior waist, bilateral wrists, and bilateral shanks. *Ambient:* appliance contact sensors; TI AWR2944PEVM radar. | Structured ADL tasks performed in the LARA laboratory; researcher-controlled task annotation; usability testing using the System Usability Scale. | Machine-learning classifier training; video-verified ground-truth labelling; classification performance assessed using accuracy and F1-score. |
|  | Phase 2: Free-living implementation | *Reduced deployment:* Axivity AX6 on the lower back; smartwatch; Axivity AX6 on the prosthesis; Axivity AX6 on the walking aid. | Seven-day home monitoring; Days 1–4 participant confirmation loop; Days 5–7 autonomous ADL classification; learned movement threshold for free-living deployment. | Lab-trained classifier applied to free-living data; prototype-memory personalisation; autonomous ADL classification; wrist-versus-back sensor comparison; protocol adherence metrics. |
| Energy expenditure | Phase 1: Lab calibration | *Gold standard:* COSMED metabolic cart. *Wearables:* four Axivity AX6 sensors bilaterally on the thighs and shanks; smartwatch for heart rate and device-based caloric estimate. *Ambient:* TI AWR2944PEVM radar for contactless activity recognition, spatiotemporal gait analysis, postural-transition detection, and exploratory vital-sign monitoring. | Treadmill walking at three speeds (1.0, 1.3, 1.5 m/s); steady-state and ramp walking protocol; seated rest between trials. | $\dot{V}O_2$ and $\dot{V}CO_2$ converted to energy expenditure (W or W/kg); regression models mapping inertial features to energy expenditure; model performance assessed using RMSE, MAE, ICC, and LoA. |
|  | Phase 2: Free-living implementation | *Shared with ADL Phase 2:* two Axivity AX6 sensors on the intact thigh and shank; smartwatch for heart rate and device-based caloric estimate. | Same seven-day free-living monitoring period; no calorimetry performed outside the laboratory. | Lab-calibrated energy-expenditure model applied to free-living data; energy expenditure estimated per activity type using ADL-linked segments; smartwatch estimates used as a plausibility reference only. |
| Emotional states | Phase 1: UK validation | *Instrument development:* custom smartwatch app; valence–arousal questionnaires; emoji-based response interface. *Ambient:* radar-based monitoring of vital signs (heart rate, respiration) as exploratory physiological inputs. | Emoji framework validation in a UK sample; valence–arousal mapping of emoji responses; PPI-informed prompt calibration. | Questionnaire-based anchoring of emoji ratings; emoji clustering and instrument reduction; cross-cultural comparison with Japanese reference data. |
|  | Phase 2: Free-living implementation | *Shared with ADL Phase 2:* smartwatch with custom EMA app. | Interval-contingent EMA prompts (four per day) during the seven-day monitoring period; participant-initiated emotional-state reports supported at any time. | Valence–arousal distribution analysis; co-occurrence analysis with ADL and energy-expenditure estimates; protocol adherence and response-completion metrics. |

Phase 2 is operationally unified across all three studies, sharing the same seven-day free-living monitoring period. ADL, activity of daily living; EMA, ecological momentary assessment; ICC, intraclass correlation coefficient; LARA, Living and Robotic Assistive Laboratory; LoA, limits of agreement; MAE, mean absolute error; PPI, patient and public involvement; RMSE, root mean square error.

The free-living implementation phase is operationally unified across all three studies: participants undergo a single seven-day home monitoring period during which all sensing modalities are deployed simultaneously. This duration was selected to capture both weekday and weekend activity patterns while balancing ecological representativeness with participant burden, consistent with established free-living monitoring frameworks [39]. The three studies are analytically distinct but logistically concurrent, each utilising the data streams relevant to its objectives.

#### Participants

The protocol is designed to include adults with unilateral lower-limb amputation who are current users of a prosthetic device. A total of approximately 40 participants will be recruited, comprising 15–20 individuals with transfemoral amputation and 15–20 individuals with transtibial amputation. For the ADL study, the target sample of approximately 40 participants is intended to capture inter-participant variability across transfemoral and transtibial prosthesis users. Model evaluation will use participant-wise cross-validation so that data from an individual participant are not shared between training and test folds, reducing similarity-related bias in activity-recognition evaluation [40]. For the energy expenditure study, each participant contributes steady-state observations at three walking speeds, yielding approximately 120 speed-referenced observations across the sample; given the low-dimensional feature set of the validated estimation pipeline [41]. For the emotional states study, the target sample supports an exploratory UK replication of the valence–arousal mapping procedure and a preliminary cross-cultural comparison with the Japanese reference data [42]. The sample additionally supports stratified observation across amputation level, enabling exploratory comparison between transfemoral and transtibial subgroups.

Inclusion and exclusion criteria were developed in accordance with criteria applied in comparable prosthetic monitoring studies [12, 43].

##### Inclusion criteria

Participants will be eligible if they: are aged 18 years or above; have a unilateral transfemoral or transtibial amputation; are current users of a lower-limb prosthesis for ambulation; are able to walk independently with or without a walking aid; and are able to provide informed consent.

##### Exclusion criteria

Participants will be excluded if they present with: an acute or unstable medical condition likely to affect mobility or safe participation during the study period; severe neurological, musculoskeletal, cardiovascular, or respiratory conditions that would prevent safe completion of the walking or activity-based assessments, as determined by the research team or supervising clinician; cognitive impairment limiting the ability to follow study procedures or provide informed consent (Mini-Mental State Examination score below 24, where clinically indicated [44]); residual-limb complications or prosthetic socket problems preventing safe prosthesis use during testing; or inability to safely complete the laboratory-based walking assessments.

### Instrumentation

Instrumentation is organised into three categories: wearable sensing systems, ambient sensing modalities, and reference systems used for validation. This structure enables systematic evaluation of sensor contribution, complementarity, and scalability within a unified protocol. An overview of all devices, sensing modalities, and technical characteristics is provided in Table 3 and Fig 1.

**Table 3.** Technical specifications of sensing devices used across laboratory and free-living phases.

| Feature | Xsens MTw Awinda | Axivity AX6 | Apple Watch Series 8 | TI AWR2944PEVM |
| --- | --- | --- | --- | --- |
| Sampling frequency | Internal: $\sim 1000$ Hz; output: 60–120 Hz | Internal: 12.5–1600 Hz; output: 100 Hz | Output: $\sim 50$–100 Hz | ADC sampling up to 45 MSps; chirp and frame rate configurable |
| Sensor resolution | N/A | 16-bit accelerometer and gyroscope | N/A | 16-bit ADC |
| Measurement range | Acc: $\pm 160$ m/s<sup>2</sup> ($\sim 16$ g); gyro: $\pm 2000^\circ$/s | Acc: $\pm 2/4/8/16$ g; gyro: $\pm 125$–$2000^\circ$/s | Acc: model-dependent; gyro: $\pm 2000^\circ$/s | 76–81 GHz FMCW radar; range, velocity and angular coverage configuration-dependent |
| Raw data access | Raw and processed data via SDK | Fully raw data stored in memory | Partially processed | Raw ADC capture supported via DCA1000EVM; processed radar point-cloud output available |
| Calibration | Advanced Kalman filter fusion (XKF); drift correction | Minimal built-in calibration; user responsible for preprocessing | Fully automatic; proprietary fusion | Built-in RF calibration |
| Synchronisation | High precision ($< 10$ μs between sensors) | Standalone recording; no built-in multi-sensor synchronisation | Limited; single-device timestamping | Internally synchronised chirp/frame timing; external synchronisation required for multi-sensor |
| Deployment | Laboratory ADL only | Laboratory and free-living | Laboratory and free-living | Laboratory and free-living |

ADC, analogue-to-digital converter; FMCW, frequency-modulated continuous-wave; IMU, inertial measurement unit; RF, radio-frequency; SDK, software development kit; XKF, Xsens Kalman filter.

**Fig 1.**
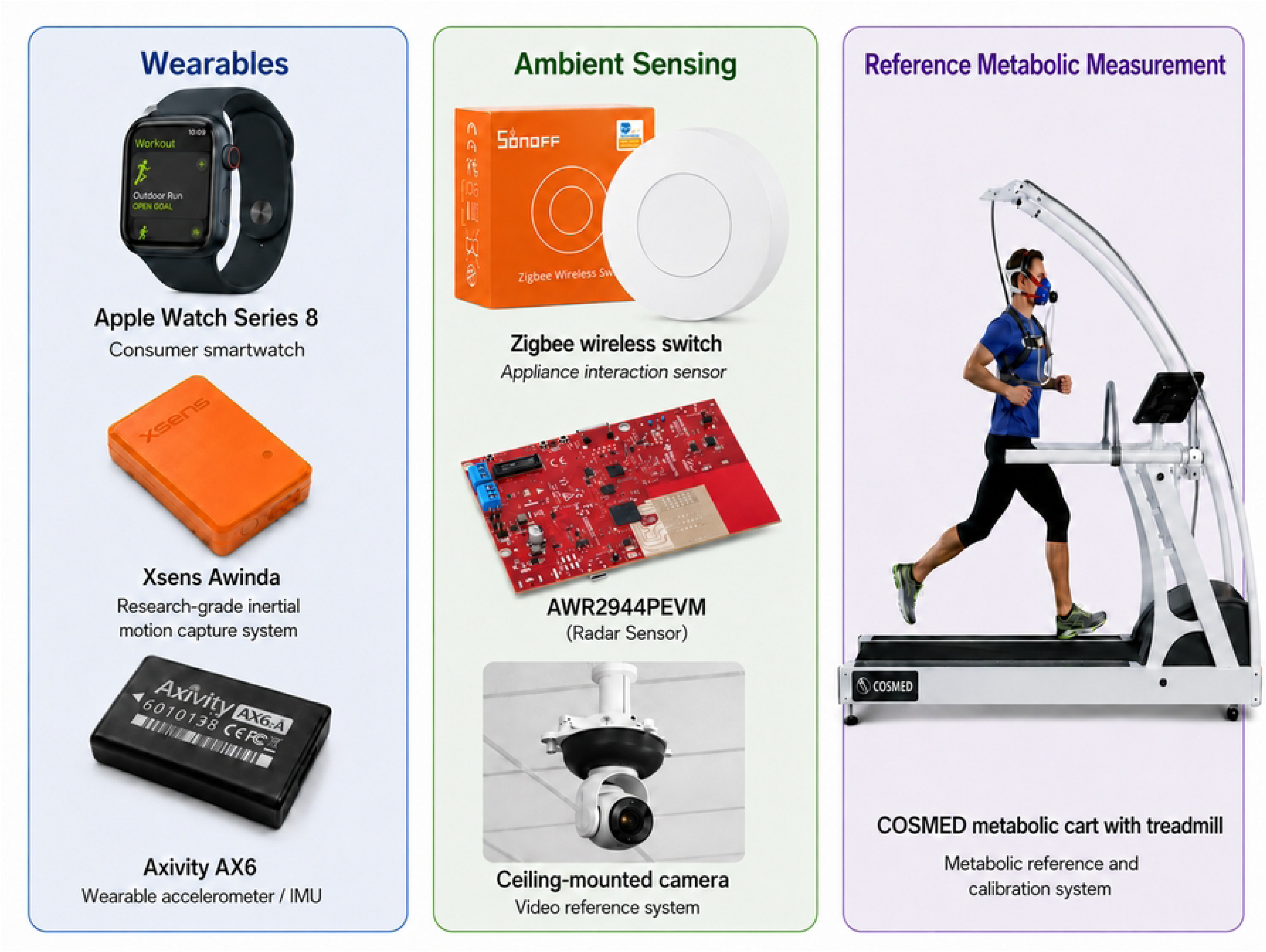
Devices used across the protocol.

#### Wearable sensing

##### Smartwatch

A wrist-worn consumer smartwatch (Apple Watch Series 8; Fig 1) is used across both laboratory and free-living phases of the protocol, serving three distinct roles. First, during laboratory-based ADL classification and validation, the device supports researcher-controlled activity annotation through the ADL Recorder, a custom application that enables real-time labelling of activity onset and offset, synchronised with concurrent wearable data streams (Fig 3). Second, the wrist-mounted inertial measurement unit (IMU; triaxial acceleration and angular velocity) provides motion data used to train and deploy the ADL classification model. During free-living monitoring, the Apple Watch (Series 8; Apple Inc., Cupertino, CA, USA) runs the trained classifier serving two functions: during Days 1–4, it prompts participants to confirm detected self-initiated activities, enabling human-in-the-loop ecological validation and prototype memory updates; during Days 5–7, the classifier operates autonomously using a learned movement threshold derived from laboratory data, requiring no participant interaction. Third, the device hosts the custom emotional state application, which captures momentary emotional states through a valence–arousal emoji interface during both laboratory and free-living phases. The Apple Watch additionally records heart rate and device-estimated caloric expenditure [45], providing complementary physiological data to support wearable-derived energy expenditure estimation during free-living monitoring [46, 47].

##### Axivity AX6 inertial sensor

The Axivity AX6 (Axivity Ltd., Newcastle upon Tyne, UK; Fig 1) is an inertial measurement unit incorporating a triaxial accelerometer (*±*2/4/8/16 g) and a triaxial gyroscope (125–2000*^◦^*/s), together providing six degrees-of-freedom inertial measurement of linear acceleration and angular velocity. The device is deployed at multiple body locations across the protocol, each serving a distinct analytical purpose.

At the lower back, a single device is worn continuously throughout the free-living monitoring period, providing whole-body motion data for offline ADL classification and characterisation. This placement is selected for its proximity to the centre of mass and its well-documented use as a sensor location for free-living mobility assessment across multiple clinical populations, including those with high gait asymmetry [48], and its acceptability during extended monitoring periods [49]. One device is additionally placed on the prosthetic device and one on the walking aid where applicable, providing contextual information on prosthesis donning and doffing, patterns of prosthesis use, and reliance on walking aids during daily activities, building on accelerometry-based classification of prosthetic use validated in transtibial users [50] and extended here to transfemoral participants [15]. These three sensors collectively support the ADL classification study.

For the energy expenditure study, four devices are worn bilaterally at the thigh and shank during laboratory calibration. This configuration adapts the thigh–shank inertial sensing approach of Slade et al., who used IMUs on the thigh and shank of one leg in healthy adults [41]; bilateral sensing is added here to characterise side-specific signals in lower-limb prosthesis users. For transfemoral participants, the prosthetic thigh position is replaced by the most proximal available location on the residual limb, documented in the session metadata. Bilateral placement enables development of a body-side-independent energy expenditure model using intact limb inertial signals as the primary input [51]. During free-living monitoring, two devices are worn at the thigh and shank of the intact limb, to which the laboratory-calibrated model is subsequently applied. Energy expenditure estimation is restricted to walking segments identified by the ADL classifier, ensuring the model is applied only to conditions consistent with its laboratory calibration.

Concurrent data collection across all body locations additionally enables offline comparison of sensing configurations, evaluating whether the lower back sensor alone can approximate the full lower-limb configuration; a key question for future sensor reduction [12, 15].

##### Xsens MTw Awinda inertial measurement units

Xsens MTw Awinda inertial measurement units (Xsens Technologies B.V., The Netherlands; Fig 1) are used exclusively during laboratory-based ADL assessment, where they serve as a high-fidelity wearable reference for whole-body movement. Sensors are positioned across the upper limbs, lower limbs, and trunk (sternum, anterior waist, bilateral wrists, and bilateral shanks) to capture distributed inertial signals during structured task execution. The multi-segment configuration provides detailed characterisation of movement dynamics under controlled conditions and supports interpretation of data from the reduced free-living sensing configuration. Xsens IMUs are not deployed during the energy expenditure protocol or during free-living monitoring, as the multi-sensor configuration would impose a substantial participant burden and is impractical for prolonged, unsupervised use.

#### Ambient sensing

Ambient sensing modalities are incorporated during laboratory-based ADL assessment to provide complementary contextual and environmental information on task execution, participant movement and interaction with the surrounding space. These systems support annotation accuracy and cross-modal verification of activity boundaries, while also contributing additional ambient cues that may support activity classification and gait analysis. In addition, selected ambient sensing outputs may provide exploratory information relevant to physiological monitoring, including vital-sign assessment where appropriate. Together, the ambient modalities complement wearable sensors and reference systems by providing supporting information for activity recognition, movement characterisation and multimodal validation during structured laboratory tasks.

##### Appliance-mounted sensors

Selected household appliances and objects within the LARA simulated home environment are instrumented with binary contact and low-power motion sensors (Fig 1). These generate event-based signals confirming object interaction during ADL execution, for example, confirming that a kettle was lifted or a cupboard door was opened. These signals complement video-based annotation by providing objective, time-stamped markers of interaction events [52].

##### Radar sensor

A short-range millimetre-wave radar platform (TI AWR2944PEVM; Fig 1) is included as an exploratory ambient sensing modality. The platform is based on the AWR2944P 76–81 GHz frequency-modulated continuous-wave (FMCW) radar sensor and provides direct connectivity to the DCA1000EVM for raw analogue-to-digital converter (ADC) data capture [53]. The device operates as an FMCW radar in the automotive millimetre-wave band and captures reflected radio-frequency signals from the participant and surrounding environment. Unlike wearable inertial sensors, the radar does not require physical attachment to the body and therefore provides a contactless and unobtrusive means of monitoring movement within the laboratory or home environment. This makes it particularly relevant for free-living assessment, where participant compliance, sensor placement, battery management, and long-term wearability can limit the reliability of body-worn systems.

Radar data can be processed to estimate the range, radial velocity and angular location of moving targets within the field of view. In the present protocol, these measurements are used to generate radar representations such as range–Doppler maps, range–angle maps, micro-Doppler signatures and point-cloud outputs. These outputs allow the position and movement of the participant to be tracked over time, supporting identification of activity transitions, locomotion periods and navigation between task locations. In contrast to binary appliance sensors, which only indicate object interaction, radar provides continuous spatial and kinematic information about the moving person. It therefore provides complementary contextual information to wearable sensors, video annotation and appliance-mounted sensors during structured ADL assessment.

The radar platform also enables extraction of movement features relevant to mobility assessment. During walking and postural transitions, radar point clouds and Doppler signatures can be used to derive spatiotemporal parameters such as walking speed, step timing, stride-related periodicity, turning behaviour, sit-to-stand and stand-to-sit transitions, and changes in movement symmetry. These measures are clinically relevant because alterations in gait speed, cadence, step regularity, turning stability and postural transition performance are associated with mobility decline and can provide early indicators of walking-affected conditions, including neurological, musculoskeletal and prosthesis-related functional changes. Accordingly, radar-derived movement features are explored as a privacy-preserving method for unobtrusive mobility monitoring and for detecting changes in functional performance over time [54, 55].

In addition to gross movement analysis, the radar can support micro-motion-based monitoring [56]. When a participant is seated, standing still or otherwise relatively stationary, small chest-wall displacements caused by respiration and cardiac activity can modulate the phase of the received radar signal. These phase variations can be analysed to estimate respiratory and heart-rate information, providing a potential contactless source of complementary physiological data [57].

Radar measurements are configuration- and environment-dependent, with the quality of the derived features influenced by sensor placement, field of view, multipath reflections, occlusion, participant orientation, and the selected chirp, frame and signal-processing parameters. Within this protocol, the radar is therefore used as a supporting and complementary sensing modality rather than as the primary source of ADL ground-truth labels. Its role is to provide an additional ambient signal for cross-modal verification, subject tracking, movement characterisation, and exploratory assessment of contactless mobility and vital-sign monitoring.

#### Reference systems for validation

##### Optical video recording

Ceiling-mounted cameras in the LARA laboratory (Fig 1) serve as the gold standard for ADL classification validation [52]. Continuous video capture of task execution enables researcher-led manual annotation of activity onset, offset, sequence, and completion. These annotations provide the ground truth labels used to train and evaluate the ADL classification model. Video recordings are used exclusively for annotation and validation purposes and are not subjected to quantitative signal processing.

##### Indirect calorimetry

A laboratory-based metabolic cart (COSMED, Italy; Fig 1) serves as the gold standard for energy expenditure estimation [41, 58]. The system is interfaced with a motorised treadmill and measures the rate of oxygen consumption (*V̇* O_2_) and carbon dioxide production (*V̇* CO_2_) on a breath-by-breath basis, enabling accurate quantification of metabolic demand during controlled walking. Prior to each session, calibration is performed following manufacturer guidelines, including flow calibration using a 3-litre syringe and gas calibration using certified reference gas mixtures. Steady-state values are derived by averaging minutes 2–5 of the six-minute steady-state walking period, excluding the first and final minutes to avoid contamination from the ramp-up and ramp-down transitions, providing the physiological reference against which wearable-derived energy expenditure estimates are calibrated [58].

### Procedures and analyses

#### ADL classification

Data collection for ADL classification is conducted in the Living and Robotic Assistive (LARA) Laboratory at Heriot-Watt University (Edinburgh, UK; LARA testbed: https://care.hw.ac.uk/projects/testbeds/LARA.html), a sensor-enabled simulated home environment incorporating kitchen, living, and transitional spaces. This setting enables systematic observation of daily activities under controlled yet ecologically representative conditions, preserving the environmental contexts in which prosthesis use typically occurs.

Participants will complete a structured set of nine ADL tasks selected to represent functional demands commonly encountered during everyday prosthesis use, including postural transitions, locomotion, load carrying, object interaction, and personal care activities (Table 4). Tasks were identified through a combination of constructs represented in established prosthetic outcome instruments (Table 1) and domains highlighted as relevant during PPI consultation; the origin of each activity is indicated in Table 4. Activities are performed at a self-selected pace, and participants are permitted to use walking aids to preserve natural movement behaviour.

**Table 4.** ADL tasks included in the laboratory protocol, their functional domain, relevance to prosthesis use, and source of task selection.

| ADL | Functional domain | Relevance to prosthesis use | Source |
| --- | --- | --- | --- |
| Sitting | Postural control | Static stability and load distribution | CI |
| Standing | Postural control | Balance and weight-bearing symmetry | CI |
| Sit-to-stand & stand-to-sit | Transfers | Weight transfer, turning, and lower-limb control | CI |
| Walking (level ground) | Locomotion | Gait symmetry, coordination, stability, and continuous dynamic balance control | CI |
| Carrying shopping bags | Load carrying | Dual-task walking and load adaptation | PPI |
| Opening a door | Object interaction | Coordination during upper-limb interaction while standing | PPI |
| Shelving items | Object interaction / coordination | Reach, balance, and upper–lower limb coordination | PPI |
| Mopping the floor | Household activity | Whole-body coordination and dynamic balance | CI |
| Brushing teeth | Personal care | Functional independence and sustained standing control | CI |

CI, clinical instrument; PPI, patient and public involvement.

The wearable sensor configuration used during this laboratory session is shown in Fig 2.

**Fig 2.**
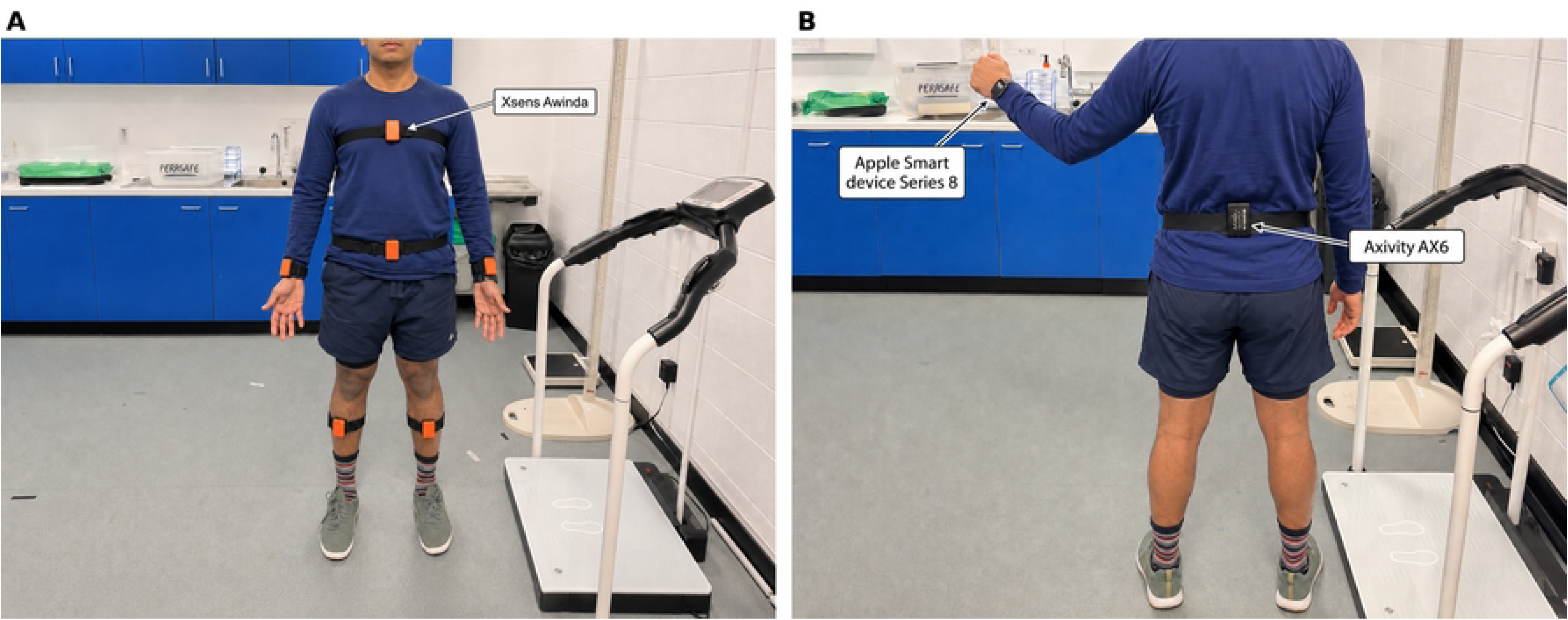
Wearable sensor configuration for the ADL classification study (Phase 1, laboratory). A: Front view: Xsens MTw Awinda IMUs positioned at the sternum, anterior waist, bilateral wrists, and bilateral shanks. B: Rear view: Axivity AX6 positioned at the lower back (L4/L5), with the Apple Watch Series 8 worn on the left wrist.

All sensing modalities operate concurrently during task execution. Activity onset and offset are annotated in real time using the ADL Recorder, a custom smartwatch and smartphone application (Fig 3). The researcher selects the activity being performed from a predefined list on the smartphone interface, triggering synchronised inertial data collection from the smartwatch and generating a timestamped label for each activity segment. This researcher-controlled annotation approach minimises reliance on retrospective segmentation and provides the primary synchronisation layer across wearable, ambient, and video data streams. Ceiling-mounted video cameras provide continuous behavioural reference to verify annotation accuracy, while appliance-mounted sensors and radar systems contribute complementary contextual information on task execution, participant movement and environmental interaction. In addition to supporting cross-modal confirmation of activity boundaries, the radar system provides an exploratory contactless sensing modality for activity recognition, gait analysis and vital-sign extraction. Radar-derived motion features may support the identification of activity transitions and locomotor periods, while movement signatures can be examined for spatiotemporal gait characteristics. During periods of limited body motion, radar data may also be explored for respiratory and cardiac activity estimation. The usability of the sensing configuration and data collection procedures will additionally be assessed in collaboration with the study’s clinical team using the System Usability Scale [59].

**Fig 3.**
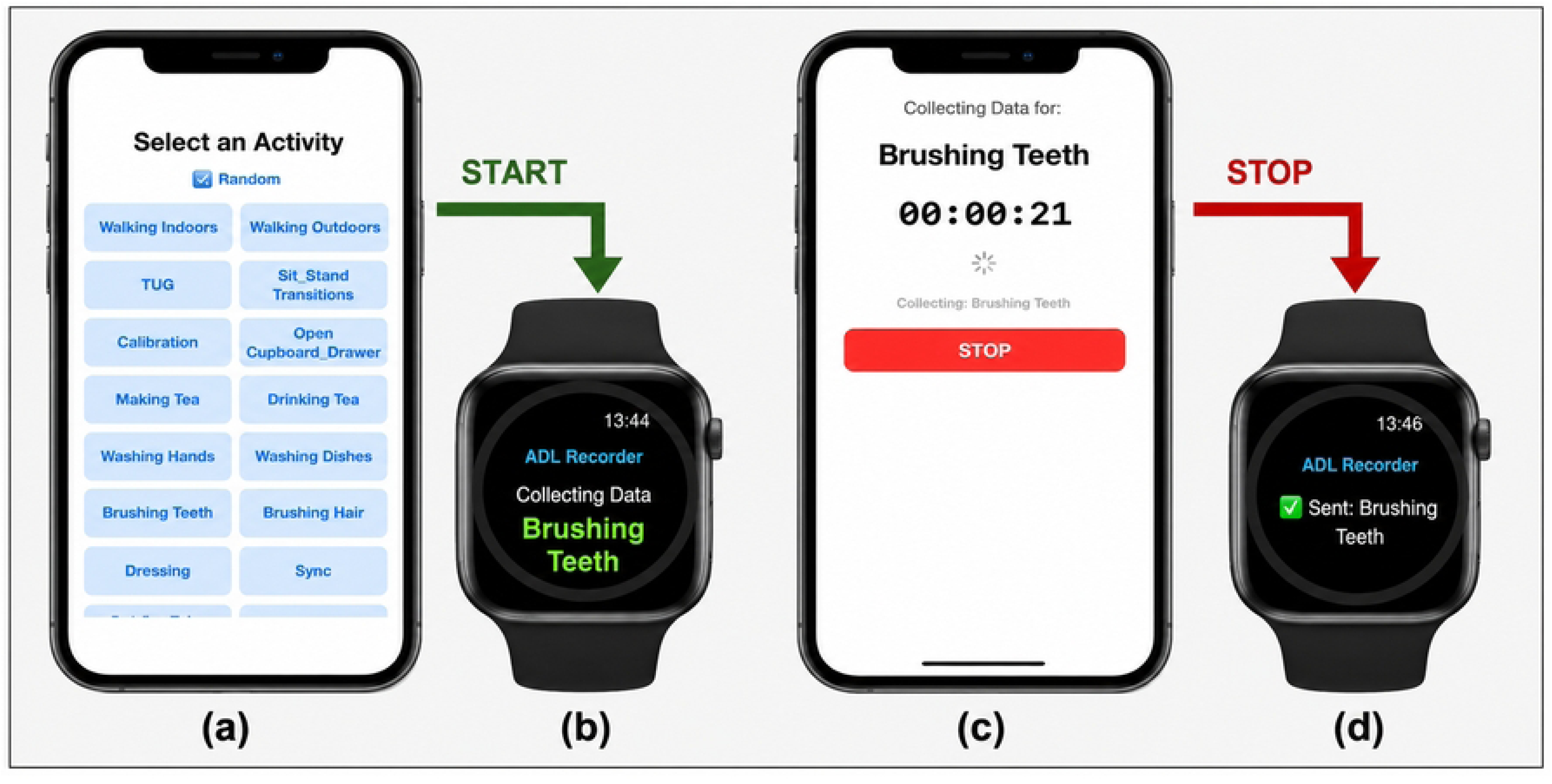
Researcher-controlled activity annotation and synchronisation interface (ADL Recorder). A: Smartphone interface displaying the predefined list of ADL tasks with randomised sequencing. B: Apple Watch confirmation of active data collection during task execution. C: Smartphone timing interface showing task duration during recording. D: Apple Watch confirmation of successful data transfer with the associated ADL label.

The ADL classification model is trained on wrist IMU data from the smartwatch, using video-verified activity labels as ground truth. Inertial signals are segmented using a sliding-window approach, with window length and overlap optimised empirically according to activity duration and classification performance, consistent with established wearable-HAR segmentation practice [60]. Signals are processed through a classification architecture combining convolutional feature extraction with temporal modelling, enabling capture of both local motion patterns and sequential activity dynamics [61]. A motion complexity score derived from signal variability and intensity is incorporated to modulate model confidence during training and inference [20]. A lightweight personalisation mechanism based on prototype memory allows class representations to be updated using confirmed activity segments without full model retraining, enabling adaptation to individual movement patterns while preserving generalisability learned from laboratory data [62]. Model performance is evaluated using participant-wise cross-validation, summarised using accuracy and F1-score. Building on validated algorithms, digital mobility outcomes will be extracted from free-living inertial data to characterise the duration, frequency, and intensity of functional activities, following the validation framework established for digital mobility outcomes [39, 48, 63].

During the free-living monitoring period, ADL classification operates across two concurrent streams. In the first stream, participant-initiated recordings during Days 1–4, participants use the smartwatch application to initiate recordings when performing recognisable activities. Upon completion of each recording, the model generates an activity prediction and presents the participant with a confirmation prompt. Confirmed labels are saved alongside the inertial data; where the prediction is incorrect, participants select the correct activity from a predefined list. This human-in-the-loop confirmation mechanism provides ecologically valid ground-truth labels in free-living, enabling out-of-lab model refinement through prototype memory updates [62] and generating corrective data that directly captures where and how model performance degrades outside controlled conditions.

In the second stream, autonomous classification during Days 5–7, the trained classifier operates without participant interaction, using a learned movement threshold derived from laboratory data to trigger classification. This stream enables unsupervised characterisation of daily activity structure, including activity type, frequency, duration, and movement complexity. These outputs are further examined in relation to prosthesis use and non-use, reliance on walking aids, and circadian patterns of activity participation across the monitoring period.

Concurrent data from sensors positioned on the prosthetic device and walking aid provide complementary contextual layers for interpreting these activity outputs. The prosthesis-mounted sensor enables detection of donning and doffing events and periods of prosthesis non-use, supporting characterisation of daily prosthesis wear patterns and identification of compensatory mobility strategies during doffed periods [15, 50]. The walking aid sensor enables detection of aid use during specific activities and across the day, providing objective evidence of reliance on assistive devices during different functional tasks. Together, these three sensing layers (activity type, prosthesis use status, and walking aid engagement) support a multi-dimensional characterisation of real-world functional behaviour that directly corresponds to the clinical outcome domains identified in Table 1.

Activity patterns are further examined in relation to time of day, enabling investigation of circadian patterns and variability in activity participation and potential fatigue-related changes in prosthesis and walking aid use across the monitoring period. Where GPS data are available from the smartwatch, they provide supplementary information on community ambulation and outdoor activity periods. Weather data extracted from publicly accessible meteorological sources are additionally linked to activity patterns to support interpretation of day-to-day variability in outdoor activity.

Offline analysis further compares classification performance between the wrist-based and back-based sensing configurations, using the confirmation-loop labels as a shared reference. This comparison evaluates the technical viability of reduced sensing configurations and informs the potential for cross-modal signal reconstruction approaches for deployable single-device systems in subsequent work.

#### Energy expenditure estimation

The energy expenditure estimation protocol adopts and adapts a previously validated treadmill-based framework in which lower-limb inertial sensing is used to estimate the energetic cost of walking [41]. The protocol is specified here for lower-limb prosthetic users, for whom energetic cost is elevated and gait variability is increased compared with individuals without lower-limb amputation [51]. Recent work has demonstrated the feasibility of estimating metabolic energy expenditure from short-duration walking tests in individuals with lower-limb amputation, supporting the extension of treadmill-calibrated approaches to this population [64].

Participants complete treadmill walking trials at three predefined speeds (1.0, 1.3, and 1.5 m/s), corresponding to slow, comfortable, and fast walking respectively, and selected to capture the range of common daily-life walking speeds observed in both non-amputees and lower-limb prosthesis users [41, 51]. Each trial consists of a 60-second ramp-up phase, a six-minute steady-state walking period, and a 60-second ramp-down phase, with seated rest periods between trials. The COSMED metabolic cart records breath-by-breath *V̇* O_2_ and *V̇* CO_2_ concurrently with lower-limb inertial data from four Axivity AX6 sensors positioned bilaterally at the thigh and shank (Fig 4). Bilateral placement enables development of a body-side-independent energy expenditure model applicable regardless of prosthesis side. The Apple Watch records heart rate and device-estimated caloric expenditure throughout each trial as supplementary physiological signals [47].

**Fig 4.**
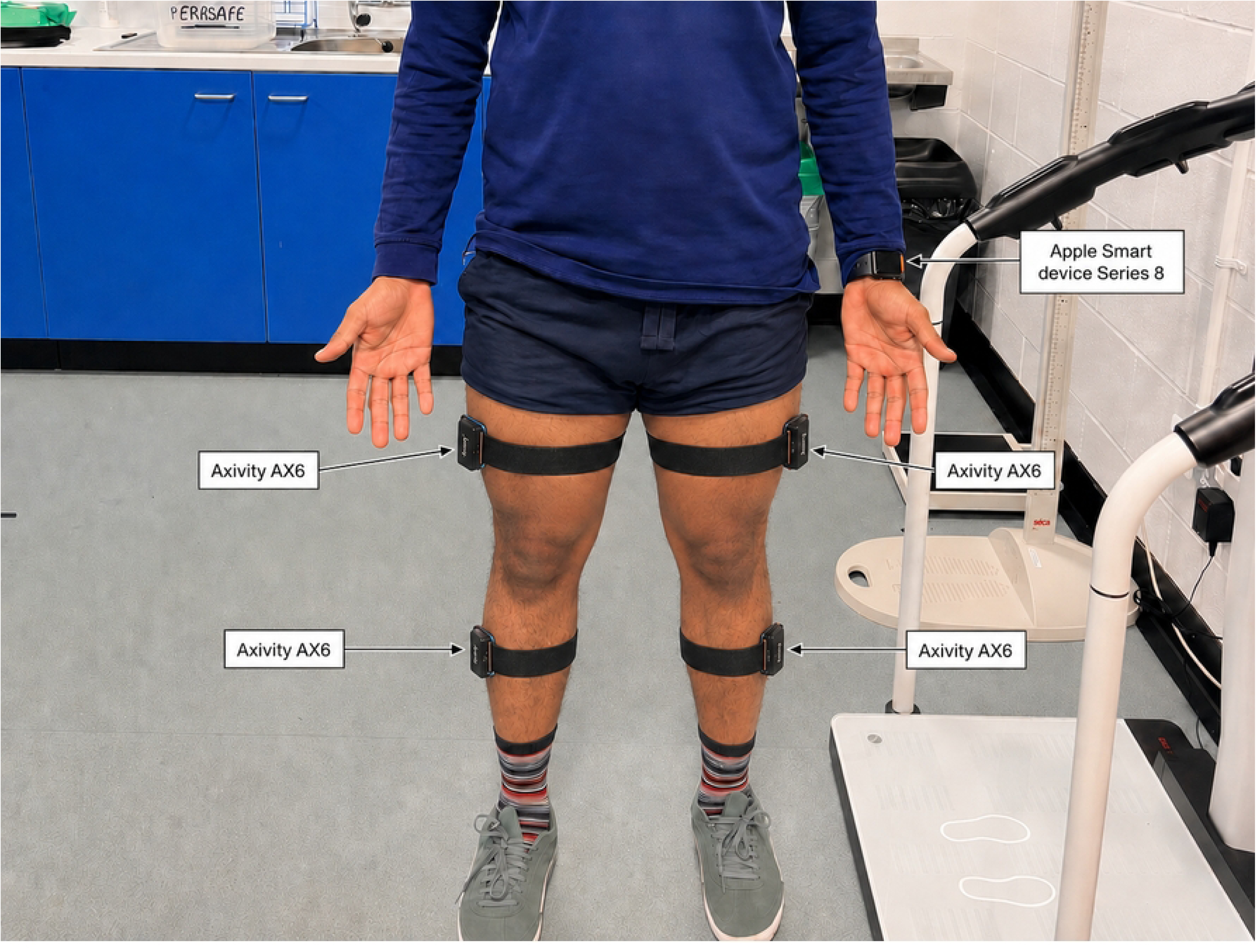
Wearable sensor configuration for the energy expenditure estimation study (Phase 1, laboratory). Four Axivity AX6 sensors are positioned bilaterally at the lateral mid-thigh and lateral mid-shank, with the Apple Watch Series 8 worn on the left wrist.

Energy expenditure is derived from steady-state *V̇* O_2_ and *V̇* CO_2_ values computed by averaging minutes 2–5 of the steady-state walking period. Concurrent inertial signals are processed using the validated pipeline of Slade et al. [41], which derives gait cycle features internally and maps them to energy expenditure via a pre-trained regression model. Participant-specific parameters such as body mass and height are optionally incorporated to improve model accuracy [41, 58]. Model performance will be evaluated using participant-wise cross-validation and reported using root mean square error (RMSE), mean absolute error (MAE), relative error, intraclass correlation coefficients, and limits of agreement [41, 65].

In free-living, the laboratory-calibrated regression model is applied offline to continuous inertial data recorded from two Axivity AX6 sensors at the thigh and shank of the intact limb. Energy expenditure estimation is restricted to walking segments identified by the ADL classifier, ensuring the model is applied only to conditions consistent with its laboratory calibration. Energy expenditure is characterised per activity type, with estimates linked to activity labels generated concurrently by the ADL classifier, enabling identification of the most energetically demanding activities during real-world prosthesis use. Smartwatch-derived caloric estimates provide a secondary plausibility reference only, given the substantial errors associated with commercial wearables in energy expenditure estimation [41, 46]. Portable calorimetry is not used in free-living owing to the practical constraints of mask-based setups and gas calibration requirements. Free-living energy expenditure outputs are therefore interpreted descriptively, with uncertainty bounds inherited from the laboratory validation metrics. Free-living walking in prosthesis users frequently comprises short, intermittent bouts that may fall below the speeds sampled during treadmill calibration; estimation is therefore restricted to identified walking segments and interpreted with this constraint in mind, consistent with evidence that metabolic cost can be estimated from short-duration walking in this population [64]. Protocol adherence outcomes including sensor wear time and data completeness are reported alongside the energy expenditure characterisation.

#### Emotional states

The emotional states component of the protocol employs a custom smartwatch application designed to capture momentary self-reports of emotional experience in free-living environments through an emoji-based valence–arousal interface. Prior to deployment, the emoji set presented to participants is derived through a structured validation process building on an equivalent study conducted in Japan [42].

The validation addresses a key practical challenge: the full emoji set comprises 74 items [42], which is impractical to deploy in full during momentary assessment. UK participants rate each of the 74 emojis on valence and arousal dimensions using the Self-Assessment Manikin [66], with emotional states conceptualised within the valence–arousal space of the Circumplex Model of Affect [67], replicating the methodology established in Japan [42]. Ratings are used to construct a valence–arousal map of the full emoji space, from which clusters of emotionally similar emojis are identified. Representative emojis are selected from each cluster to yield a reduced set that spans the full valence–arousal space while remaining practical for momentary self-report. This process enables direct cross-cultural comparison of emoji perception between UK and Japanese populations [42]. Prompt frequency and format are additionally calibrated based on PPI feedback to ensure that the sampling protocol is acceptable and minimally disruptive during everyday life.

The reduced emoji set is embedded within the custom smartwatch application. In free-living, the waking day is divided into four equal time blocks determined individually based on each participant’s reported wake and sleep times. Within each block, a single prompt is delivered at a randomly selected time, yielding four assessments per day over the seven-day monitoring period (maximum 28 prompted assessments per participant) [68]. At each prompt, participants select the emoji that best represents their current emotional state. Participant-initiated reports are also supported, enabling capture of emotional responses to specific activities or events outside the scheduled prompt schedule. Concurrently, the application records heart rate and heart rate variability, providing objective physiological context alongside subjective emotional reports. Where GPS data are available from the smartwatch, they provide supplementary contextual information on community ambulation and outdoor activity. All responses and concurrent data streams are time-stamped and synchronised with the wearable inertial data, enabling examination of emotional fluctuations in relation to inferred ADL types and estimated energy expenditure.

Analysis focuses on the distribution of valence–arousal states over time, temporal variability in emotional experience, and co-occurrence with activity type identified by the ADL classifier and energy expenditure estimated from the inertial regression model. These outputs are interpreted descriptively and exploratorily, with no causal assumptions made regarding the relationship between emotional state, physical activity, or energy expenditure. Protocol adherence outcomes including prompt adherence, response completeness, and participant-reported acceptability are reported alongside the emotional state characterisation.

### Operationalisation

#### Data standardisation and multimodal harmonisation

Following established approaches for multimodal data standardisation in digital health research [69], reliable integration of data acquired from heterogeneous sensing systems requires explicit procedures for synchronisation, structuring, and storage. As illustrated in Fig 5, all sensor outputs are organised using a standardised hierarchical folder structure in which raw files are preserved and systematically grouped by study, sensing modality, and data type. Sensor outputs are stored as time-stamped files in non-proprietary formats, with one file per device and sensor location. Accompanying metadata files document sensor configurations, acquisition parameters, device placement, and contextual information associated with each recording session.

**Fig 5.**
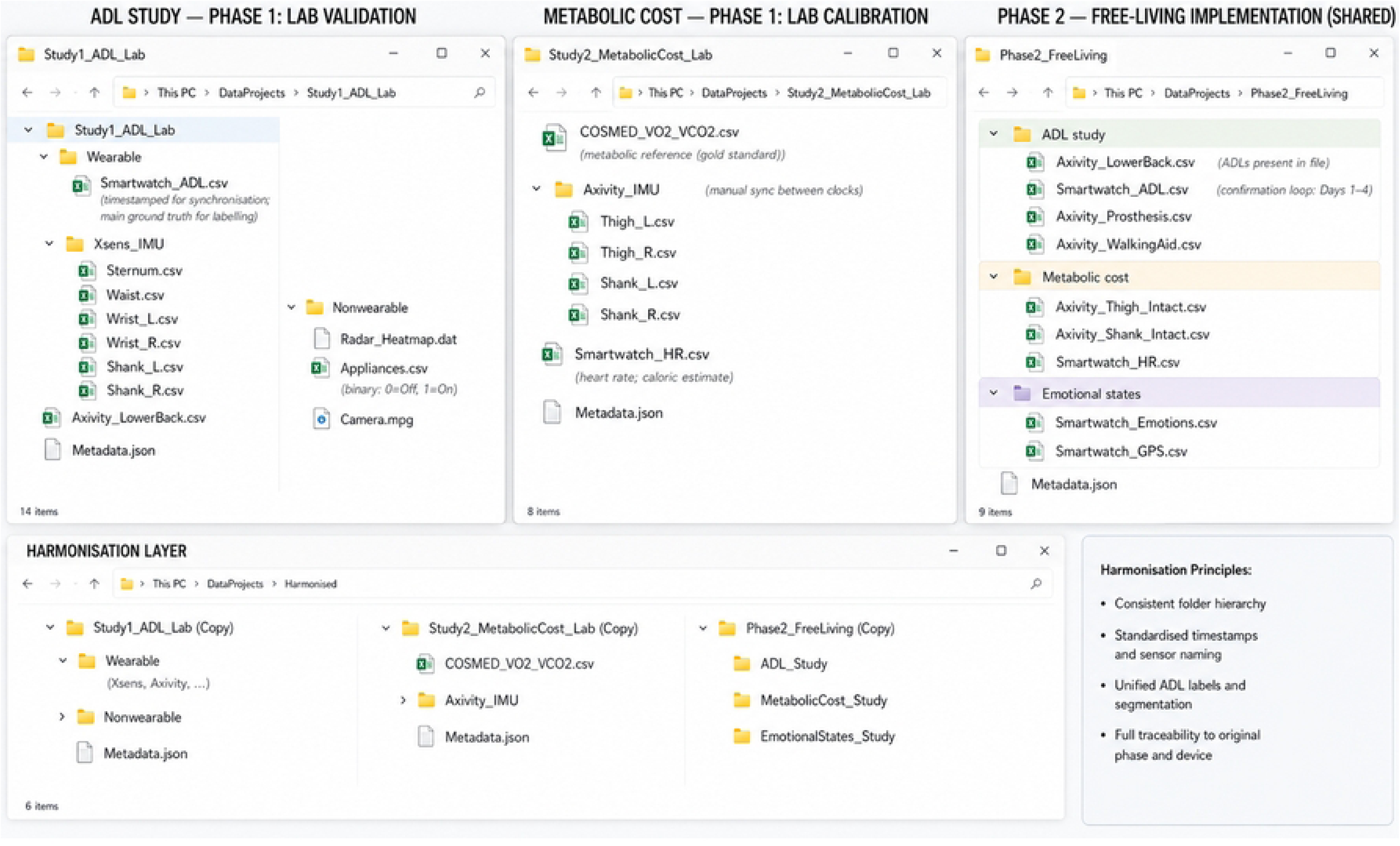
Standardised folder hierarchy for raw sensor data, grouped by study, modality, and device, with accompanying metadata. A harmonisation layer applies uniform naming conventions, timestamps, and sensor identifiers across laboratory and free-living datasets while preserving traceability to original sources.

Temporal harmonisation is performed explicitly at the data level through post hoc alignment of synchronisation events across recording systems. During laboratory-based assessments, synchronisation events are executed while all recording systems are active, generating identifiable temporal markers across wearable, physiological, ambient, and video streams. These markers are used during post-processing to align device clocks and correct for offsets or drift, with synchronisation information preserved in associated metadata files.

During free-living monitoring, where explicit synchronisation events are not feasible, temporal consistency is maintained through calibrated device timestamps and protocol-defined acquisition procedures. Timestamp integrity is verified during harmonisation using a dedicated synchronisation pipeline implemented in Python, which standardises timestamps across devices, corrects for clock offsets, and aligns data streams using shared signal features where explicit markers are unavailable. Signals are resampled to a common temporal resolution and consistency checks are performed to identify temporal drift, missing segments, or misalignment.

Harmonisation does not involve merging or modifying raw sensor files. Instead, it establishes a common structural and temporal representation that preserves links to original data sources and enables reference models and calibration parameters derived under laboratory conditions to be applied consistently to free-living datasets.

#### Preprocessing and quality control

Prior to analysis, all harmonised datasets undergo structured preprocessing and quality control to ensure signal integrity, temporal consistency, and suitability for subsequent modelling, following established procedures for multimodal wearable data processing [69].

Inertial sensor signals are inspected for completeness, continuity, and plausibility based on expected dynamic ranges and sampling characteristics. Periods of signal dropout, saturation, or implausible values are detected using predefined thresholds and flagged for exclusion. Non-wear periods in free-living recordings are identified using established accelerometer-based criteria and removed prior to analysis. Where required, signals are filtered to remove high-frequency noise while preserving movement dynamics relevant to activity classification and energy expenditure estimation. Preprocessing parameters including filter characteristics and segmentation schemes are defined a priori and applied consistently across participants and devices.

Following harmonisation, temporal alignment is verified by locating shared synchronisation markers across data streams and confirming that event timestamps are consistent across modalities. Laboratory datasets exhibiting excessive clock drift or incomplete synchronisation are excluded from analysis according to device-specific tolerance criteria. For free-living data, timestamp continuity and the presence of expected movement and rest patterns are used to verify temporal integrity.

Physiological signals from indirect calorimetry are screened for calibration errors, abnormal respiratory values (respiratory exchange ratio outside the range 0.7–1.0), and non-steady-state behaviour (coefficient of variation of *V̇* O_2_ exceeding 10% in the final two minutes of each trial), with analysis restricted to segments meeting predefined stability criteria [41, 65]. Video and ambient reference data are used solely to verify timing and completeness of laboratory tasks and are not subjected to quantitative signal processing. Ambient sensing data are screened for completeness, timestamp consistency and expected event structure. Appliance-mounted sensors are used to confirm object interaction events, whereas radar data undergo additional signal processing to support exploratory contactless movement analysis.

Radar preprocessing includes removal of static clutter, generation of range–Doppler and range–angle representations, and detection of the moving subject within the radar field of view. Detected radar points are then associated across successive frames to support subject tracking and to characterise changes in position and movement over time. Where appropriate, Doppler signatures are further analysed to extract micro-Doppler information associated with limb and trunk motion during walking, postural transitions and other ADL tasks. These radar-derived outputs provide complementary features for activity recognition, gait analysis and cross-modal verification of task execution.

#### Ethical considerations

Ethical approval for all studies described in this protocol was granted by the School of Engineering and Physical Sciences Research Ethics Committee at Heriot-Watt University (reference 2025-12121-16250). The study is conducted in accordance with the Declaration of Helsinki. All participants provide written informed consent before taking part, and may withdraw at any time without giving a reason.

#### Safety considerations

The physical demands of the protocol are concentrated in the laboratory-based walking and activity assessments. Treadmill trials are supervised throughout, with a clinician available, and include seated rest periods between trials. Walking speeds are individually capped at a level the participant can sustain safely, and any trial may be stopped at the participant’s request or by the supervising researcher. The exclusion criteria screen out individuals for whom the assessments would pose an unacceptable risk. Wearable devices used during free-living monitoring are non-invasive. Adverse events are recorded and reported to the approving ethics committee.

#### Data management

Raw sensor data are stored on secure, access-controlled institutional infrastructure and managed in accordance with Heriot-Watt University’s research data management policy. Video recordings and other potentially identifying information are stored separately from sensor data and accessible only to authorised members of the research team. Data shared with collaborating institutions for analysis are de-identified before transfer. Each recording is accompanied by metadata documenting sensor configuration, placement, and acquisition parameters.

#### Study status and timeline

Ethical approval has been obtained and the study is in progress at the time of submission; no results are yet available and no data have been analysed. Recruitment commenced in May 2026 and is anticipated to conclude in January 2027. Data collection, comprising the laboratory assessments and the seven-day free-living monitoring period for each enrolled participant, is conducted on a rolling basis and is anticipated to be completed by March 2027. Data analysis will commence thereafter, with results anticipated by December 2027. No participant has completed the full protocol at the time of submission.

## Discussion

This study presents a structured multimodal protocol for assessing lower-limb prosthesis use in free-living environments by integrating measures of functional activity, metabolic cost, and emotional state within a unified and validated framework. The primary contribution lies in establishing a multimodal, multi-dimensional methodology that aligns wearable-derived measures with clinically relevant constructs commonly assessed in prosthetic outcome instruments.

### PPI co-design elements

Patient and public involvement contributed substantively to the design and feasibility of the proposed protocol. Both prosthetic users and clinicians emphasised that functional activity, metabolic effort, and lived experience represent complementary dimensions of prosthesis use that cannot be adequately captured through a single modality, consistent with emerging perspectives on stakeholder-informed design in digital health systems [70].

PPI input directly informed several key protocol decisions. Feedback from prosthetic users shaped the selection of ADL tasks, leading to the inclusion of load carrying, household interaction, and object manipulation tasks that reflect genuine daily demands, and the exclusion of stair negotiation as less representative of typical home environments. Clinicians highlighted the importance of capturing movement at the level of the prosthetic limb and assistive devices, directly informing sensor placement on the prosthesis and walking aids, consistent with clinician perspectives on the role of activity monitoring in prosthetic service provision [71]. Concerns regarding monitoring burden and usability led to a streamlined sensor configuration and a seven-day monitoring duration, balancing ecological representativeness with participant acceptability.

Feedback on emotional self-report confirmed its relevance as a contextual dimension of prosthesis use. Prosthetic users consistently noted that externally observed functional performance does not always capture the complete, contextualised, or variable nature of their experiences during short assessments. Participants described pain, frustration, and fatigue that may not be apparent during task execution, yet substantially influence everyday prosthesis use [18, 19]. Clinicians acknowledged the importance of these dimensions, while noting that their interpretation requires careful consideration given the influence of factors beyond prosthesis use on momentary emotional state. These considerations informed both the inclusion of the emoji-based valence–arousal interface and the calibration of prompt frequency to minimise disruption to daily routines.

Data governance and trust were consistently identified as determinants of acceptability. Prosthetic users emphasised that willingness to participate in unsupervised monitoring depended on confidence in how data would be managed, protected, and used. These findings informed the incorporation of clear data management and communication procedures into the protocol and reinforce broader calls for transparency and participant agency in digital health research.

### Functional activity classification

The emphasis on activity-level classification reflects recognised limitations of aggregate mobility metrics such as step count, which provide limited insight into task-specific demands, context, or participation, and may be sensitive to device placement and proprietary processing algorithms [23]. Even when monitoring extends beyond the clinic, activity is often reduced to volume-based metrics that fail to reflect meaningful functional behaviour [12]. By focusing on classification of discrete ADLs, the proposed framework supports the interpretation of movement as functional behaviour that is directly comparable to clinical outcome constructs and more representative of prosthesis use in free-living environments [13].

Beyond discrete activity classification, the unsupervised stream enables characterisation of daily functional behaviour across multiple dimensions. Activity duration, frequency, and sequencing provide quantitative indices of participation and functional independence, while biomechanical outputs including movement complexity, gait symmetry, and compensatory strategies offer insight into how activities are performed rather than simply whether they occur [24, 48]. These outputs are examined in relation to prosthesis and walking aid use to characterise how assistive device use patterns shape daily activity participation [15, 50]. Circadian analysis of activity distributions across the monitoring period provides insight into how mobility and fatigue fluctuate throughout the day [12]. Where GPS data are available from the smartwatch, activity patterns are contextualised according to environmental domain, distinguishing outdoor community ambulation from indoor domestic activity [72]. These multidimensional outputs extend the scope of prosthetic outcome assessment beyond what is achievable with conventional clinical instruments alone [12, 13].

The two-stream free-living architecture, combining continuous passive classification with participant-initiated ecological validation, addresses a fundamental challenge in activity recognition research: the absence of ground truth labels in unsupervised settings [25]. The human-in-the-loop confirmation mechanism generates ecologically valid labelled data in free-living conditions, enabling out-of-laboratory validation of the classifier and providing correction data that reveals where and how model performance degrades outside controlled conditions. This represents an innovative methodological contribution that extends beyond the specific application to prosthetic users.

### Energy expenditure estimation

Metabolic cost represents a critical but underrepresented dimension of prosthesis use, related to the physiological demands of ambulation [51, 73]. Traditional assessments often rely on steady-state treadmill protocols conducted under controlled conditions, which do not reflect the intermittent and task-oriented nature of daily activity [51]. Moreover, traditional laboratory-based treadmill assessments may fail to capture differences in perceived fatigue and usability reported by prosthesis users in everyday life [74], as controlled steady-state conditions do not replicate the intermittent nature of real-world ambulation or the influence of factors such as poor prosthetic fit, component misalignment, and design limitations [27].

In this framework, indirect calorimetry establishes laboratory-based reference relationships with wearable data that are subsequently applied in free-living conditions, enabling metabolic cost to be interpreted in relation to specific ADLs. This approach provides insight into how effort accumulates and influences participation across the day, directly addressing constructs of functional sustainability that are rarely captured by existing outcome instruments.

### Psychosocial experience

Psychosocial experience is incorporated through ecological momentary assessment (EMA) [68] aligned temporally with activity and physiological data. This approach addresses limitations of retrospective questionnaires, which are often detached from the context in which prostheses are used. Emotional and psychological factors play a central role in prosthesis acceptance, satisfaction, and quality of life [75], and long-term user engagement with device design in everyday life [76]. By capturing emotional states in real time and mapping them with wearable-derived measures, the framework enables contextual interpretation of lived experience alongside functional and physiological outputs.

The derivation of a reduced, representative emoji set through valence–arousal clustering addresses a practical challenge in ecological momentary assessment: balancing instrument sensitivity with participant burden. The replication of the Japanese validation methodology with a UK population enables cross-cultural comparison of emoji perception and interpretation, while contributing a reusable and validated instrument for future EMA research in rehabilitation contexts.

### Multimodal integration

A key methodological contribution of this framework lies in the integration of multimodal sensing at the level of study design, rather than relying on post-hoc data fusion during analysis. Multimodal approaches are frequently proposed to overcome the limitations of single-sensor systems, yet integration is often implemented only at the analytical stage, leading to fragmented datasets and ambiguous interpretation [24]. In contrast, the present framework assigns specific predefined roles to each sensing modality and structures data acquisition accordingly, enabling more coherent interpretation of behavioural, physiological, and psychosocial measures. This design also improves robustness in free-living conditions, where transitions, environmental variability, and heterogeneous movement patterns commonly challenge activity recognition systems [25].

Furthermore, the concurrent deployment of wrist-based and back-based inertial sensors during free-living monitoring enables systematic comparison of sensing configurations. This not only informs future sensor-reduction strategies aimed at improving usability and scalability but also supports exploration of cross-modal signal reconstruction approaches for the development of deployable single-device monitoring systems.

### Ecological validity

The protocol addresses the need for improved ecological validity in prosthetic assessment. A substantial body of existing research remains constrained to laboratory-based evaluations that reflect capacity under controlled conditions rather than performance in free-living environments [12, 13]. Even when free-living monitoring is implemented, interpretation is often limited by simplified outcome metrics or lack of contextual integration. By combining laboratory validation with structured free-living monitoring, the proposed framework provides a reproducible pathway for linking controlled measurements to prosthesis use in free-living environments. This aligns with broader system-level perspectives that emphasise the importance of integrated sensing ecosystems in the development of next-generation rehabilitation technologies [77].

## Data Availability

No datasets were generated or analysed during the current study. Upon study completion, de-identified data will be deposited in the Open Science Framework and made available, subject to the governance and consent conditions approved by the ethics committee and the data-sharing agreements between the participating institutions.

## Acknowledgments

The authors would like to thank Dr Mauro Dragone (School of Engineering and Physical Sciences, Heriot-Watt University) for his support in the design and integration of the Zigbee-based appliance-mounted sensing component of this protocol. The authors also wish to acknowledge the support provided by the National Robotarium (Edinburgh, UK) for providing facilities that facilitated the conceptual development and design of the data collection framework described in this protocol.

